# The relationship between human mobility measures and SAR-Cov-2 transmission varies by epidemic phase and urbanicity: results from the United States

**DOI:** 10.1101/2021.04.15.21255562

**Authors:** Nishant Kishore, Aimee R. Taylor, Pierre E. Jacob, Navin Vembar, Ted Cohen, Caroline O. Buckee, Nicolas A. Menzies

## Abstract

Global efforts to prevent the spread of the SARS-COV-2 pandemic in early 2020 focused on non-pharmaceutical interventions like social distancing; policies that aim to reduce transmission by changing mixing patterns between people. As countries have implemented these interventions, aggregated location data from mobile phones have become an important source of real-time information about human mobility and behavioral changes on a population level. Human activity measured using mobile phones reflects the aggregate behavior of a subset of people, and although metrics of mobility are related to contact patterns between people that spread the coronavirus, they do not provide a direct measure. In this study, we use results from a nowcasting approach from 1,396 counties across the US between January 22nd, 2020 and July 9th, 2020 to determine the effective reproductive number (R(t)) along an urban/rural gradient. For each county, we compare the time series of R(t) values with mobility proxies from mobile phone data from Camber Systems, an aggregator of mobility data from various providers in the United States. We show that the reproduction number is most strongly associated with mobility proxies for change in the travel into counties compared to baseline, but that the relationship weakens considerably after the initial 15 weeks of the epidemic, consistent with the emergence of a more complex ecosystem of local policies and behaviors including masking. Importantly, we highlight potential issues in the data generation process, representativeness and equity of access which must be addressed to allow for general use of these data in public health.

## Introduction

Global efforts to prevent the spread of the SARS-COV-2 pandemic in early 2020 focused on non-pharmaceutical interventions like social distancing; policies that aim to reduce transmission by changing mixing patterns between people ^1–3^. As countries have implemented these interventions, aggregated location data from mobile phones have become an important source of real-time information about human mobility and behavioral changes on a population level. An unprecedented amount of mobile phone data has become available as a result, and mobility metrics have not only been widely used by researchers modeling SARS-CoV-2 dynamics, but also by policymakers as an indicator of social activities that drive transmission. It remains unclear, however, whether these metrics are reliable proxies of the face-to-face contact patterns that underlie SARS-COV-2 transmission.

Human activity measured using mobile phones reflects the aggregate behavior of a subset of people, and although metrics of mobility are related to contact patterns between people that spread the coronavirus, they do not provide a direct measure. During the first lockdowns in the USA in cities like New York and Seattle, the early epicenters of transmission, large changes in mobility were observed in many different data streams from mobile operators and social media ^4^. Reductions in travel, in time spent at home, and in visits to commercial areas and stores, for example, followed policies such as declarations of emergencies, shelter-in-place orders, school closures, and the cancellation of mass gatherings ^5^. Furthermore, following this dramatic change in mobility, upticks in movement patterns and travel associated with “lockdown fatigue” and re-opening preceded an increase in confirmed cases, hospitalizations, and deaths. These near real-time data streams therefore provided a useful indicator of the societal changes that were subsequently reflected in other measures of disease burden, such as diagnoses and hospitalizations, and have been used considerably to better understand the human mobility related transmission dynamics of the pandemic.

However, the multiple sources of mobility data—for example from Facebook, Google, mobile operators, or Ad Tech companies like Safegraph or Unacast—are aggregated in different ways by different providers, and have not been rigorously shown to reflect local mixing patterns accurately. Furthermore, the heterogeneous processing steps these data providers take to compute mobility metrics obscure what different proxies mean with respect to transmission. In particular, it remains unclear how or which mobility indicators to use as robust metrics for making policy decisions about SARS-COV-2 transmission in any particular location. This is critical, given the appetite of policy-makers and the general public for leading indicators of COVID-19 spread that can be used to guide personal behavior and government responses

The effective reproduction number (R(t)), together with the community incidence rate, is the most generalizable measure of the local trajectory of the epidemic because it indicates whether cases are growing, shrinking, or holding steady. But the R(t) is not always straightforward to estimate, and confirmed cases are unreliable and likely biased in different ways in different locations ^6^. Efforts in nowcasting the effective reproductive number have relied on available data and subject matter expertise on their data generation processes. In the simplest epidemiological models, the reproduction number is linearly related to the contact rate, for which mobility metrics are a proxy.

Here, we use results from a nowcasting approach from 1,396 counties across the US between January 22nd, 2020 and July 9th, 2020 to determine the R(t) along an urban/rural gradient. For each county, we compare the time series of R(t) values with mobility proxies from mobile phone data from Camber Systems, an aggregator of mobility data from various providers in the United States. We show that the reproduction number is most strongly associated with mobility proxies for change in the travel into counties compared to baseline, but that the relationship weakens considerably after the initial 15 weeks of the epidemic, consistent with the emergence of a more complex ecosystem of local policies and behaviors including masking. Importantly, we highlight potential issues in the data generation process, representativeness and equity of access which must be addressed to allow for general use of these data in public health.

## Methods

We collected data on 1,396 counties across the United States which had at least 100 cases as of June 30th, 2020. We grouped these counties into six ordinal categories defining a gradient from urban to rural as defined by the National Center for Health Statistics (NCHS) (Figure 1A) ^7^. Due to geographic heterogeneity in initial outbreaks, varying amounts of data are available by county and region of the United States (Figure 1B). In these data and for our analysis we considered NCHS category 1 (Large central metro) to be the most urban and NCHS category 6 (Non-core) to be the most rural.

**Figure 1:**
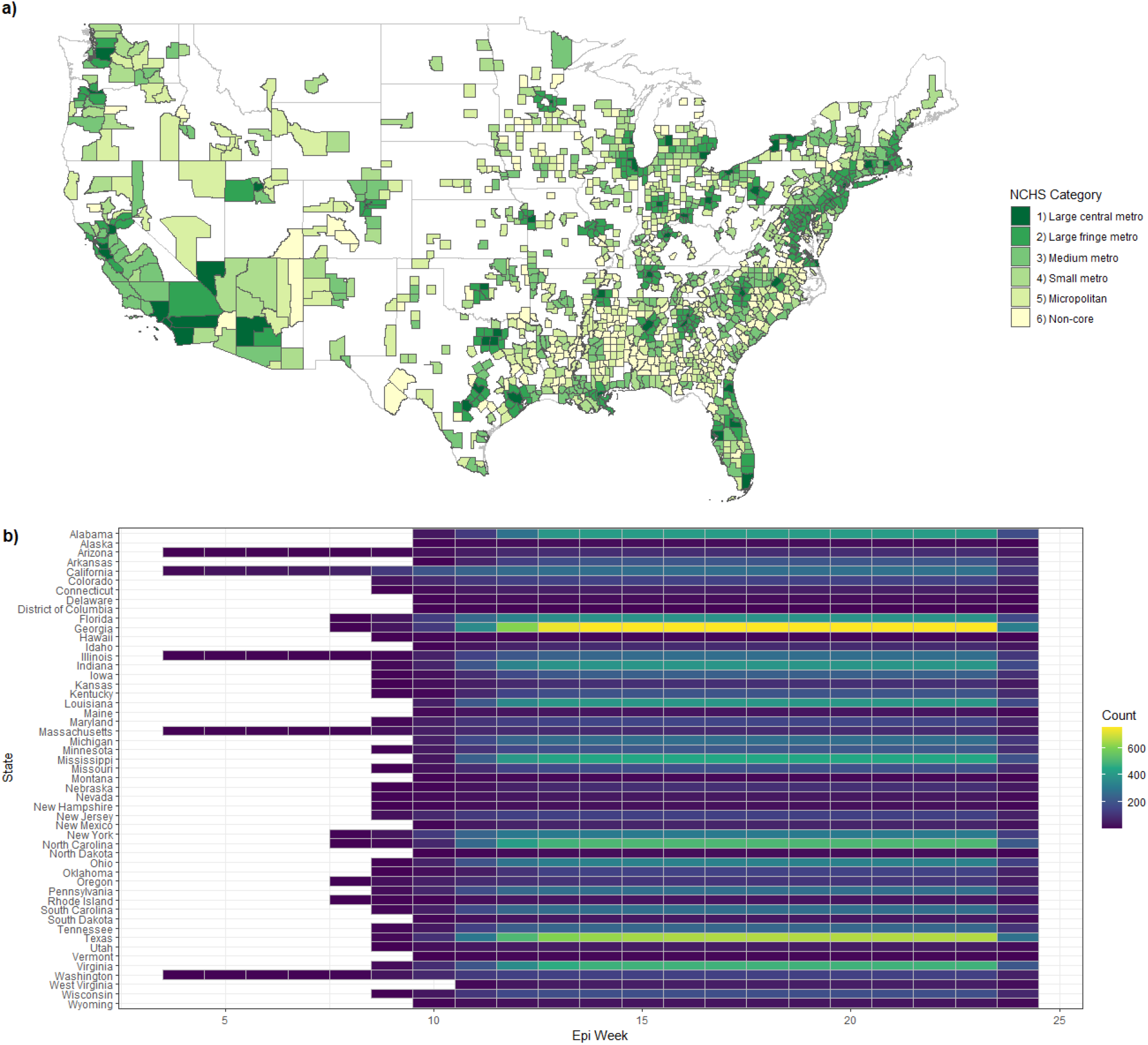
A) Counties included in the study colored by their NCHS category ranging from Large central metro (most urban) to non-core (most rural). B) The number of counties with available estimates, by state and epiweek.

For each county we collected epidemiologic data on cases and deaths, as well as human mobility metrics collated by advertisement technology (ad-tech), derived from GPS trace data provided by Camber Systems^8,9^. With the ad-tech data we calculated mobility metrics including the % change in movement into counties (% change in), % change in movement out of counties (% change out), and % change in movement within counties (% change within), as well as the average radius of gyration (RoG), average number of locations users visited (Dwell) and average entropy of movement (Entropy) for users across all counties, every day. Movement was categorized in 8 hour blocks of time with a vector of movement defined as occurring from the county where an individual spent most of their time in the preceding 8-hour block to the county where an individual spent most of their time in the current 8-hour block. We used the average movement in the month preceding the first day for which we have data as our baseline conditioning for time of day and day of the week. Descriptions of these metrics and their uses have been previously described^9^. All metrics were calculated using global position system (GPS) coordinates published by mobile phone applications for commercial purposes (ad-tech). In Figure 2 we show all metrics that were included for a county selected uniformly at random.

**Figure 2:**
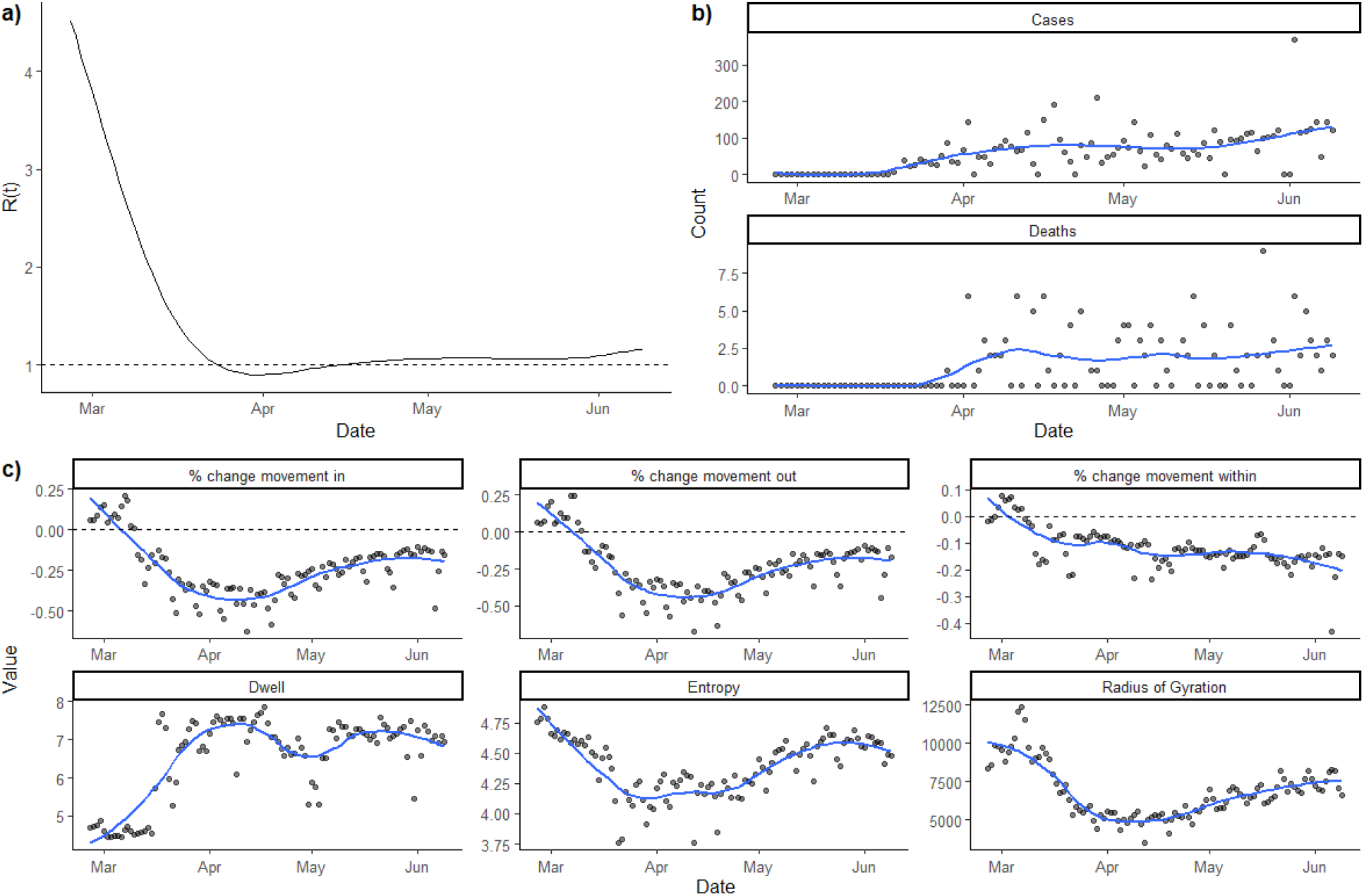
Example of metrics used for analysis for Shelby County, Tennessee which is categorized as a “Large Urban Metro” by the NCHS with a LOESS curve added.

We used daily- and county-specific estimates of the effective reproductive number generated using a bayesian nowcasting procedure which incorporated the effects of reporting delays and change in how cases were defined over time and in different regions ^10^. We aggregated all of our data points to their average log transformed value for each epidemiologic week, where epiweek is as defined by the CDC.

We specified models to predict the modelled R(t) values using the mobility data. First, we evaluated the six candidate mobility metrics and ensured that no highly correlated pairs of metrics remained in our model. Given the nature of the data generation process for both the independent variables and the outcome we used a linear model which allowed for autoregressive correlated errors. This is supported by the time-dependent structure of residuals in models where within-county correlations are not taken into account. We provide an evaluation of other model structures in our Supplementary Appendix.

We evaluated our final specification by comparing both the fit (Pearson correlation of original and predicted R(t)) and AIC. Given the difference in ad-tech data quality between rural and urban areas, we estimated the model separately for each of the NCHS county rural/urban categories. We hypothesized that the association between mobility metrics and SAR-CoV-2 may change over time and in alternate specifications included an interaction term to differentiate between the first and second half of our study period. Finally, in our last specification we also allowed the coefficients of the mobility metrics to change as a smoothed function of time to ensure that any failure of performance in our model performance would not be due to stringent model specifications. We evaluated the performance of our final model by comparing the predicted R(t) to the nowcasted R(t). Finally we evaluated the operational effectiveness of our best model at various thresholds of R(t), to determine when they could identify epidemic ‘surges’ (value of R(t) above a given threshold) using both agreement and Cohen’s kappa. We tested the robustness of our final model against both daily and weekly measures of input data.

## Results

In initial data exploration we identified that weekly “% change in” and “% change out” were very highly correlated, and so we removed % change out from our analyses (Supplemental Figure 1). We also included an interaction term between weekly average radius of gyration and entropy as, in practice, these metrics are difficult to interpret individually. Our full model, therefore, includes the estimated effective reproductive number as the dependent variable and % change in, % change within, dwell, radius of gyration, entropy and an interaction term between radius of gyration and entropy as our independent variables. We log transformed both dependent and independent variables prior to analysis.

Using the full model, we evaluated the relationship between the independent and dependent variables for subsets of the data defined by each NCHS category. As shown in Figure 3, both the fit and performance of our full model dropped off considerably in rural areas, relative to urban areas. This phenomenon persists in data that only use the data before and after week 15 (Figure 3). To avoid false confidence in model results for non-urban centers we restricted our analyses to counties in the three most urban NCHS categories (Large central metro, Large fringe metro and Medium metro). Next, we tested regression specifications containing subsets of the predictors in the full model, to identify a parsimonious set of independent variables with reasonable predictive performance.

**Figure 3:**
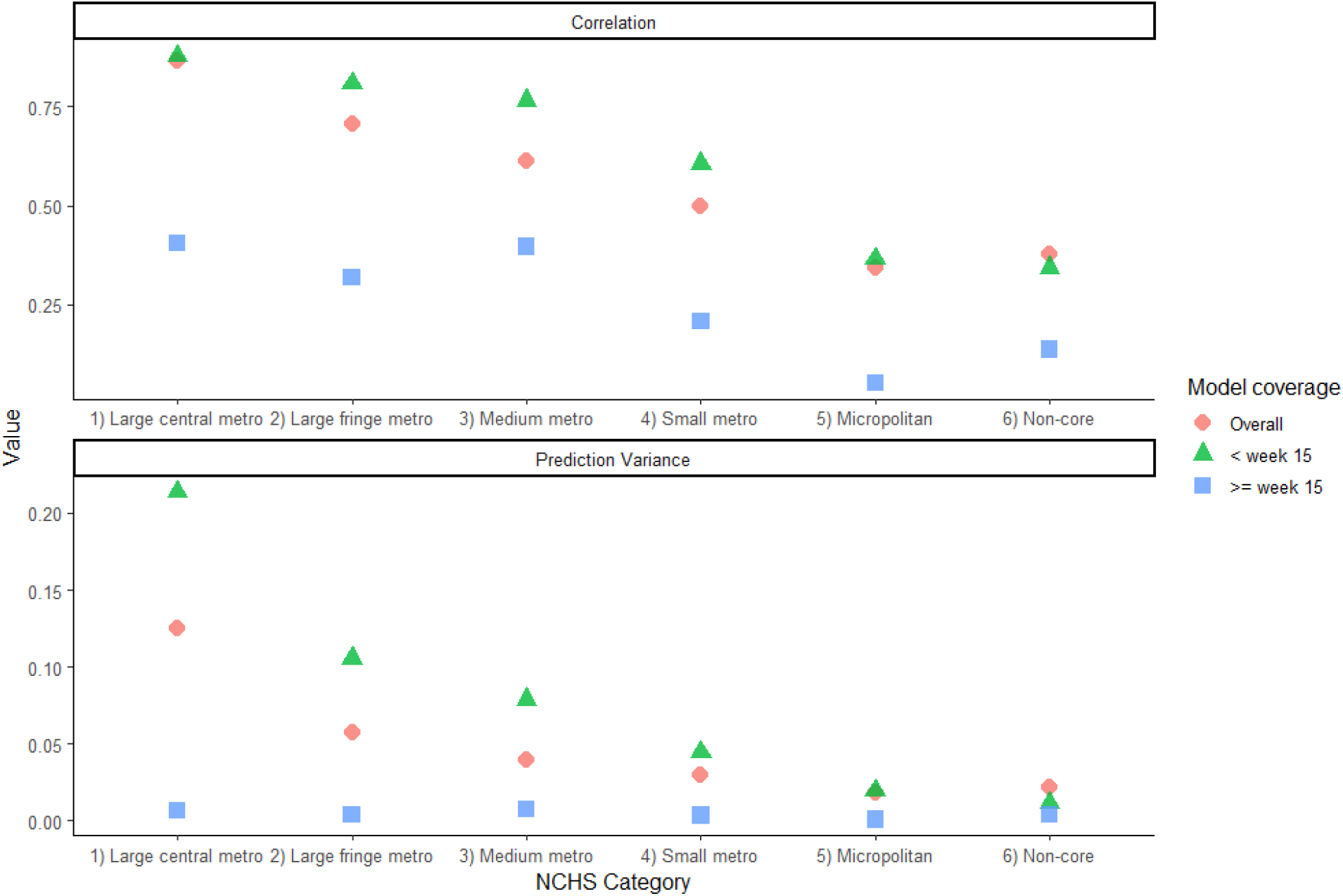
Simple correlation between nowcasted R(t) and modeled R(t) along with the prediction variance of our fully specified model for each subset of NCHS category. We show that the correlation drops drastically as we move from urban to rural areas.

**Figure 3:**
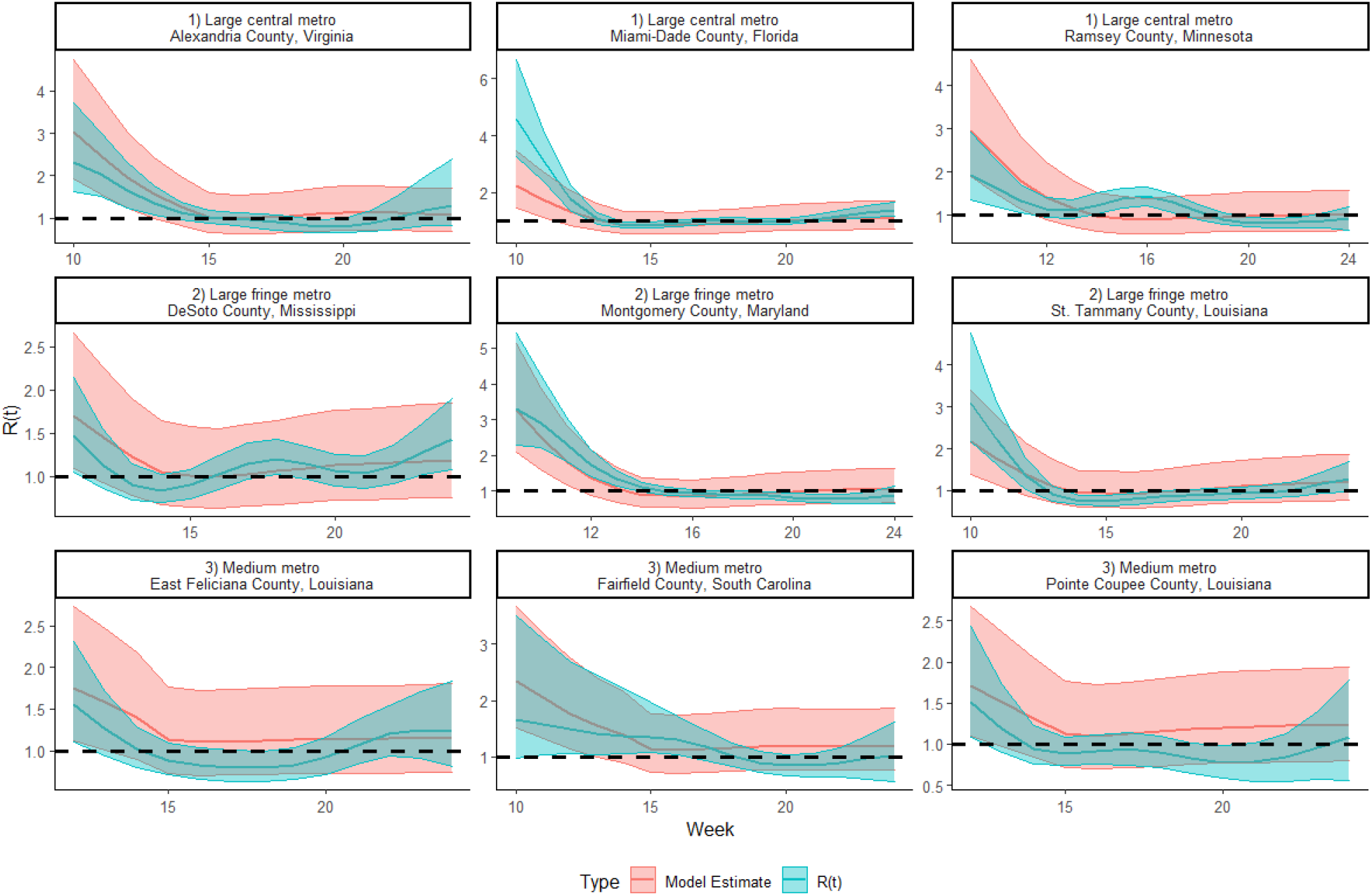
R(t) and variability estimates from the best model (red) compared to the nowcasted R(t) for three randomly selected counties in each of the three most urban NCHS categories.

We evaluated all possible combinations of independent variables, and identified the best models as those that minimize AIC. The top five models in terms of AIC are described in Table 1. The best performing model contains all variables except for radius of gyration and includes an interaction term differentiating the period of time before and after week 15. In Figure 3, we evaluate our best model in random selections of counties from each NCHS category. We found that all model predictions overlap with the estimated R(t), however, this fit changes over time, with poorer fit in later weeks. As expected, there is an increase in overall model error and decreased fit as we move from urban to rural regions.

**Table 1:** Model specifications for the top 5 models when sorted by AIC overall and when subsetting for models with only two variables along with the overall model variance (sigma).

| Model Summary | AIC | Sigma |
| --- | --- | --- |
| % change in + % change within + Dwell + Entropy + Dwell*Entropy + time_flag + % change in*time_flag + % change within*time_flag + Dwell*time_flag + Entropy*time_flag + Dwell*Entropy*time_flag | -16347.54 | 0.226 |
| % change in + % change within + Dwell + Entropy + Dwell*Entropy | -15184.3 | 0.249 |
| % change in + % change within + RoG + Dwell + Entropy + Dwell*Entropy | -15179.665 | 0.249 |
| % change in + % change within + Dwell + Entropy | -15144.628 | 0.246 |
| % change in + % change within + Dwell | -15131.36 | 0.248 |

To further evaluate the value of mobility metrics as indicators of transmission, we calculated the percent agreement between our model estimates of R(t) and the nowcasted R(t) under varying scenarios. In Figure 4 we defined agreement as either: a) agreement between our model predictions and the true value on the sign of weekly change in R(t) for a given week, and; b) agreement about whether the predicted value and the true value were both greater than 1 for that given week. In all cases our models performed well in predicting the direction of change in R(t) and if R(t) crossed 1, however, this performance declined markedly after week 14. In Figure 4 we compare these results alongside the second best model which did not include the interaction effect of time. Similarly, we evaluated the reliability of our models by calculating Cohen’s Kappa for each week, with agreement defined as the predicted value and the true value both being greater than 1 for a given week. As shown in Figure 5, we found that the fully specified model allowing for interaction with time at week 14 performs best, but with poorer performance in the second half of our study period. In all cases we saw a clear deterioration in model performance as we moved from urban to rural counties. To further investigate this deterioration in model performance we evaluated the distribution of predicted R(t) compared to the nowcasted R(t) (Figure 6). We found that the reduction in model performance in the second half of the study period is driven by the reduction in variability of the nowcasted R(t) estimates. In other words, the mobility metrics continued to change, however, R(t) did not. When plotting all the mobility metrics by county along with the log(R(t)) we find a similar result with the average log(R(t)) remaining flat after week 15 while the mobility metrics generally continued to vary (Figure 7).

**Figure 4:**
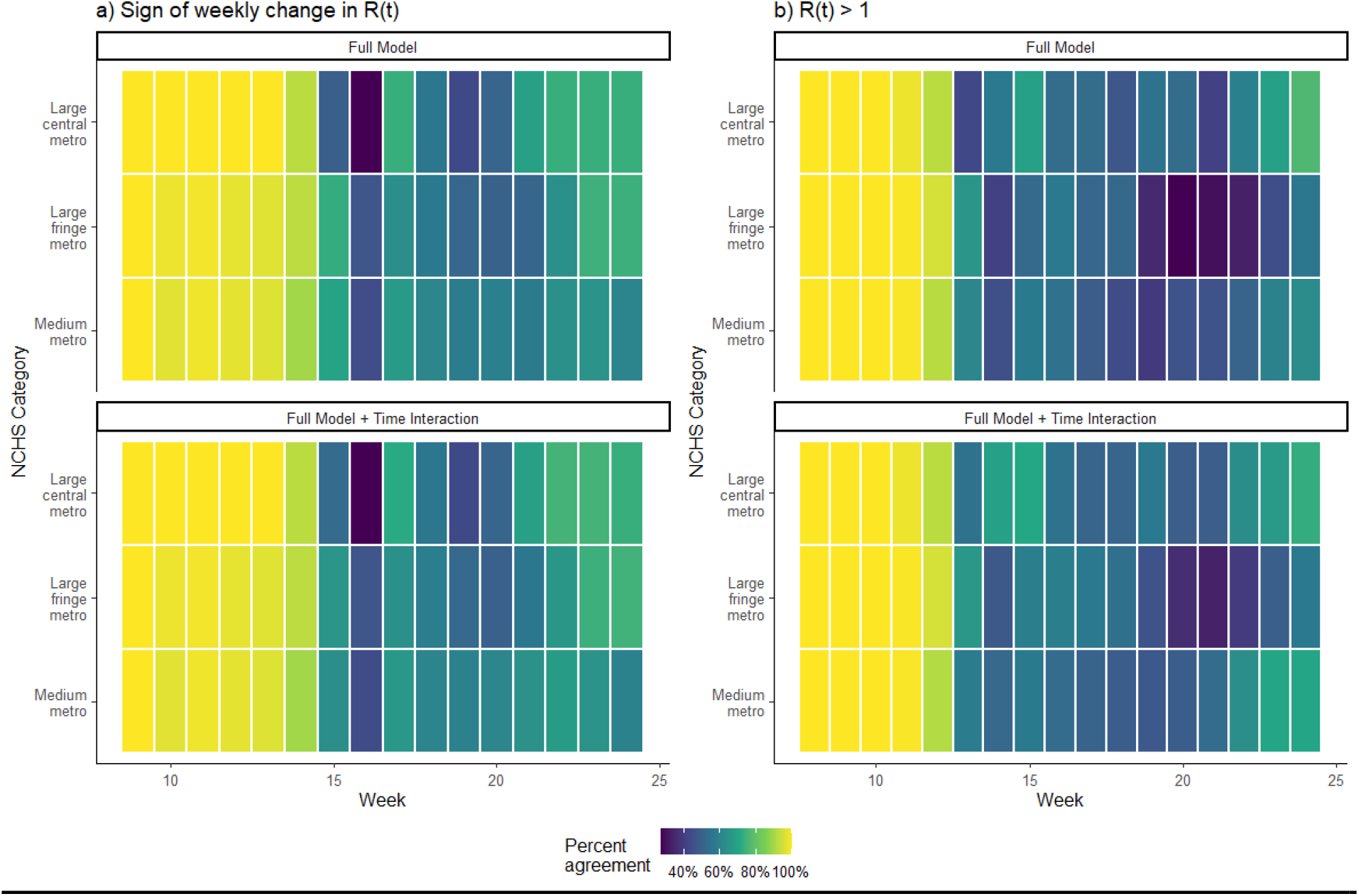
A) Percent agreement between the model estimate of R(t) and nowcasted R(t) defined as having the same sign of week on week change in R(t). B) Percent agreement between the model estimates of R(t) and nowcasted R(t) defined as predicting an R(t) above 1.

**Figure 5:**
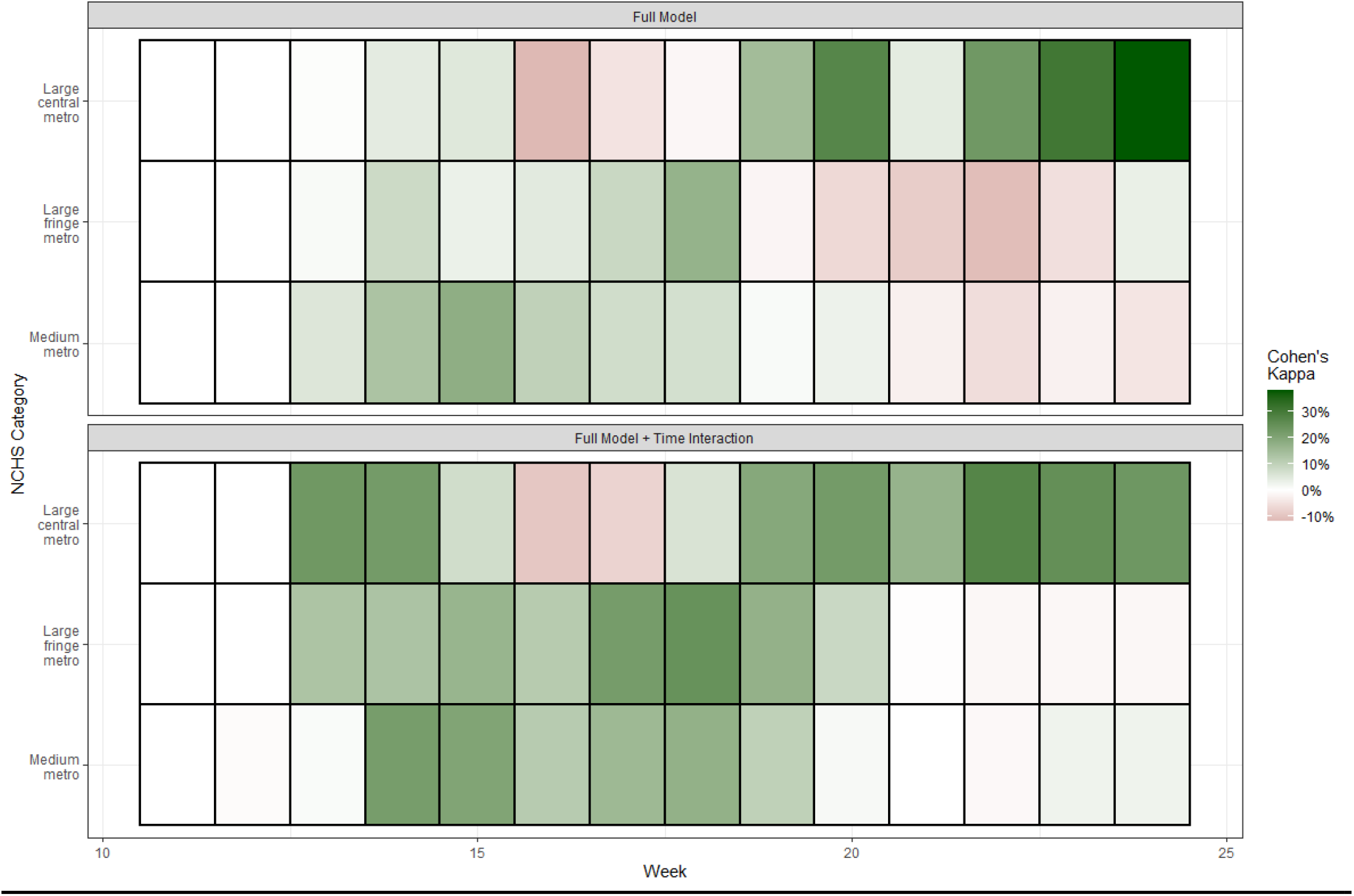
Cohen’s Kappa by week for the inter-rater agreement for having an R(t) above 1.

**Figure 6:**
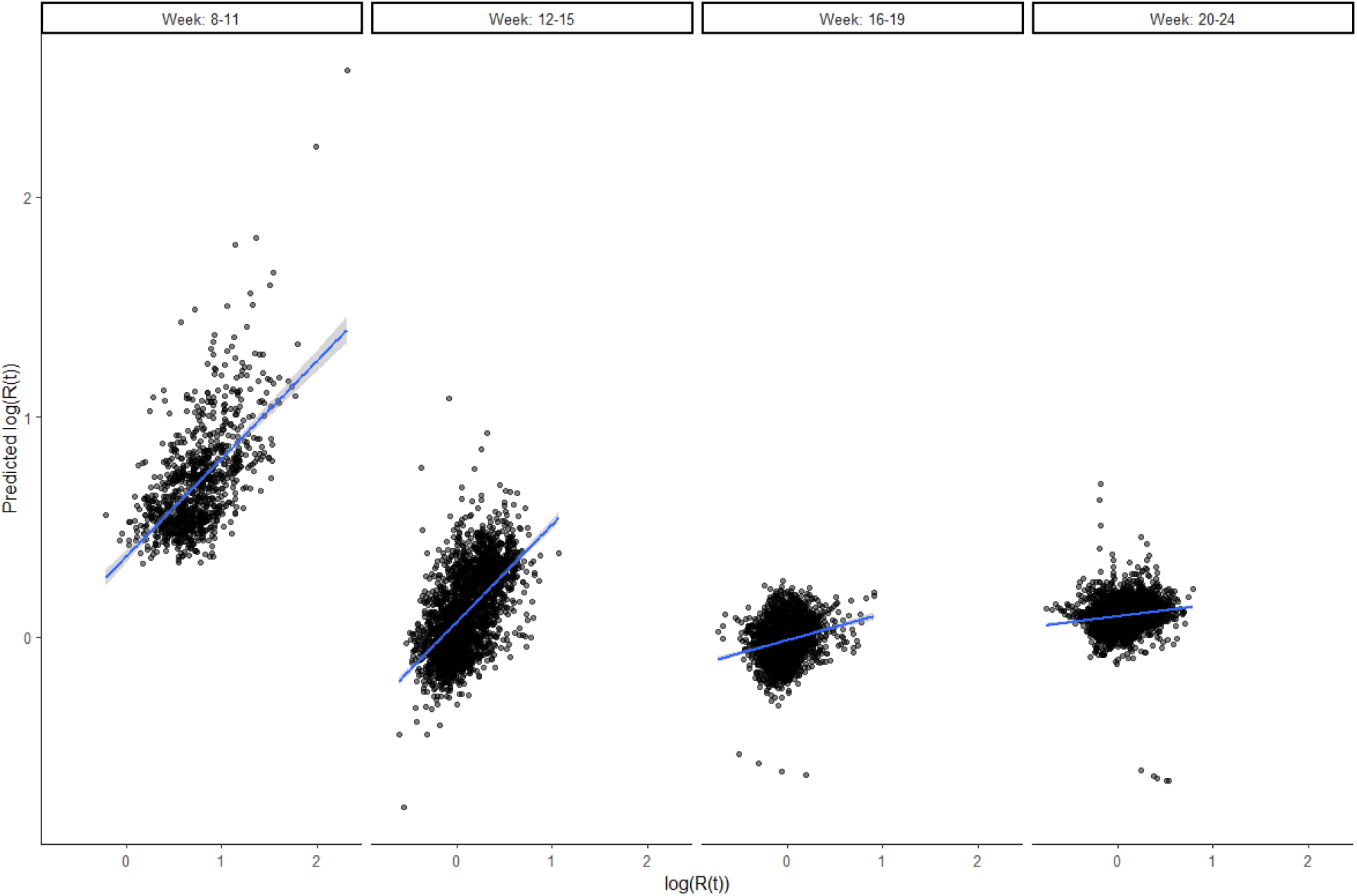
Correlation between log(R(t)) and the predicted log(R(t)) over the course of the study period. The model results are highly correlated with R(t) in the first half of the study period but performance drops off in the second half.

**Figure 7:**
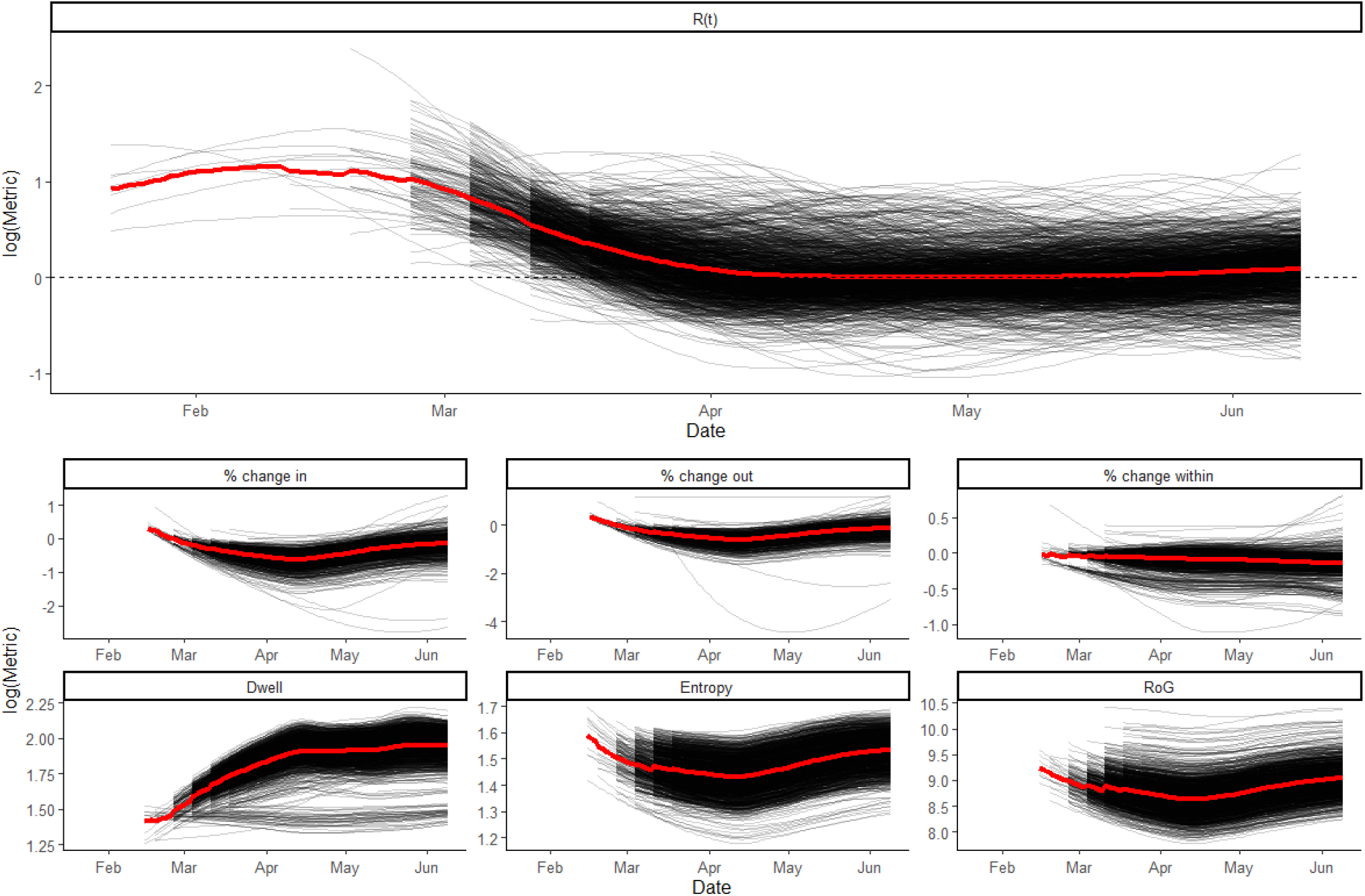
County specific plots of mobility metrics and R(t) (all on the log scale) in black with the average day specific value for each metric in red.

Allowing all predictors in the fully specified model to vary over time we found that “% change in” and “% change within” largely drive the performance of the model, with changes in covariate estimate for “% change in” coinciding with the decrease in predictive power of the model (Supplemental Figure 1).

## Discussion

Mobility data have been used extensively during the SARS-COV-2 pandemic to understand disease transmission between populations, identify hotspots of transmission and guide policy interventions. In this study we used simple and generalizable mobility metrics derived from ad-tech data to evaluate the relationship between these metrics and the effective reproductive number of SARS-COV-2 in counties in the United States during the initial period of the epidemic, from epi weeks 9 through 24 of 2020. Our model performs well for the first half of the study period with high agreement under various scenarios until the end of epi week 13. As shown in Figure 7, this reflects the period of time where, nationally, the effective reproductive number decreased from its earlier highs and showed little change through the rest of the study period. For example, in the three most urban classifications of counties, on average the log(R(t)) decreased by 0.24 in week 12 when compared to week 11. However, from week 15 onwards, the log(R(t)) didn’t change by more than 0.05 compared to the previous week. On the other hand, the ad-tech derived mobility metrics continued to change over the course of the study period.

In our final model, during the first half of our study period a 10% increase in the percent change of number of individuals traveling into a county is associated with a 5% increase in the effective reproductive number, accounting for the effects of dwell, entropy and percent change in the number of individuals moving within a county. By allowing our coefficients to vary over time we can see that this effect was larger during the first half of our study period, peaking at week 13, and diminished slightly over time (Supplemental Figure 2). These findings are useful for future use and interpretation of mobility metrics. We caution that because the relationship between the aggregated mobility patterns picked up by mobile phone data of this kind may have shifting associations with the face-to-face interactions driving transmission, these mobility metrics are unlikely to have sustained, robust predictive power for understanding R(t).

First, we noted a marked reduction in model performance in the three most rural county classifications in the United States. While this may be a function of the delayed outbreaks in these counties compared to early peaks in locations such as New York, it is important to note that the data generation process for mobility data may differ greatly between urban and rural counties. Subsequent analyses on previous studies have reported similar phenomena ^11^. For example, the types of mobile applications (apps) that individuals use, factors which lead to users sharing location information and essential trips that users take generate the raw GPS data used to calculate these metrics. These can vary immensely from county to county. This implicit and perhaps unavoidable selection of users who generate these data may also complicate the interpretability of these metrics. Furthermore, an intricate market of publishers, providers and aggregators clean, transform and extract value from these data, often making simple connections to human behavior difficult. Finally, the ad-tech pipeline is generally built for and therefore optimized for commercial purposes. This means that behaviors which may be important for public health purposes may not be those which are captured on device or enriched post-extraction. Much of this information on user demographics, coverage, app usage and post-extraction transformation is opaque, making comparisons between counties with varying urbanity or even socio-economic status difficult. These data can be potentially useful, however, further research, validation, and standardized frameworks for data generation are needed to better understand how variable processing algorithms of raw GPS traces relate to mobility patterns of the overall population, and what this means for the interpretation of mobility metrics.

Second, evaluating more parsimonious models allows us to infer some general rules of thumb regarding mobility metrics and their association with R(t) while identifying major limitations in the creation and interpretation of these metrics. Besides percent change in the number of individuals traveling into a county, entropy and dwell remain in our parsimonious model. As entropy, or the unpredictability of individuals movement increases, R(t) also increases. Anecdotally this process makes sense as individuals traveling to more locations than usual may be increasing their contact network. However, a decrease in the number of locations that a user visits can also be considered to be unpredictable and therefore result in higher entropy, underscoring the impact of appropriate baselines. Changes in entropy from week to week with no exogenous interventions should be interpreted in a different light than changes in entropy during the implementation of a travel restriction. Dwell, or the average number of locations an individual in a county spends at least 5 minutes, had an unexpected negative relationship with R(t) in our parsimonious model, likely due to artifacts of the way in which these data are collected on device. In following orders for travel restrictions or physical distancing, one might reasonably expect this metric to decrease and that individuals may travel to fewer locations. However, in our data, dwell increases almost universally in the aftermath of such interventions. This is again likely due to fewer individuals traveling by car, and instead, taking more walks with their mobile devices, and generating more locations where they spend at least 5 minutes. These issues again highlight the importance of researchers being able to interrogate the data generation process and understand the rationale behind default conditions (such as spatial limits which define a unique location, minimum number of interactions needed to generate information or the amount of time a user needs to spend in a location to register as being there) which are used to define metrics. An optimal threshold of defining locations every 5 minutes versus 10 minutes can dramatically change the metric output with no clear understanding of its impact on public health inference.

Finally, we find that the strength relationship between mobility metrics and the effective reproductive number declines dramatically over time. Allowing the coefficients of our mobility metrics to change over time did not strengthen the relationship. Nor do we consider this issue to be unique to this specific data aggregator as they consolidate sources used by a variety of other providers which, as described previously, can have opaque and overlapping user bases. There are several reasons why the relationship between mobility metrics and R(t) weakened over time. The first is exogenous universal public health measures such as masking and physical distancing. These measures intrinsically change the relationship between human movement and disease generation as contact rates between individuals may stay the same, but the probability of transmission given contact can decrease with appropriate masking and distancing. The relationship between human mobility and the effective reproductive number may also be confounded by unmeasured components of human behavior which can both drive mobility and adherence to public health measures resulting in a decrease of R(t). It is also possible that, with the smaller changes in R(t) observed after the initial epidemic response, the magnitude of signal decreased relative to the inherent noise of the mobility metrics, and estimation uncertainty consequently increased. If true, mobility metrics may be dependable indicators of gross changes in transmission behavior, but less useful for identifying smaller changes.

It is possible that a more complex model, inclusion of other variables, information on demographics or other details about human behavior may better explain the relationship between human mobility metrics and R(t). However, the non-independence of mobility data across time and locations mean that there are fewer degrees of freedom available than indicated by the massive amount of data available, and fitting flexible models to these data risks producing models with good in-sample fit but poor predictive validity. Given the opacity around the data generation process, uncertainty about the populations that these metrics represent and lack of frameworks which optimize the generation of these metrics for epidemiological inference, we chose to fit simple models, while accounting for the within-county correlation inherent in both the mobility and R(t) data.

An abundance of human mobility metrics from various providers are now available and in some cases actively integrated into both modeling and public policy efforts. These data provide information on human behavior at a level previously reserved for the data generators and their commercial interests. Our study shows that the integration of these metrics into modeling efforts can be fruitful in some cases, linking these metrics directly with public health outcomes of interest such as the effective reproductive number. In the early period of the epidemic in the United States, we infer a clear relationship between an increase in the number of individuals traveling into a county compared to a pre-pandemic baseline and an increase in the effective reproductive number. These data showed impressive potential for future public health efforts during the early stages of the SARS-CoV-2 pandemic in the United States. Nevertheless, transparency in the data generation process, investigation into representative populations, continuous revaluation of these metrics, standards for optimization of metrics for public health purposes and, perhaps most importantly, frameworks for equity of access to these data are still needed to truly unlock its potential.

## Data Availability

Data available through providers.

## Supplementary Appendix

**Supp. Fig 1.**
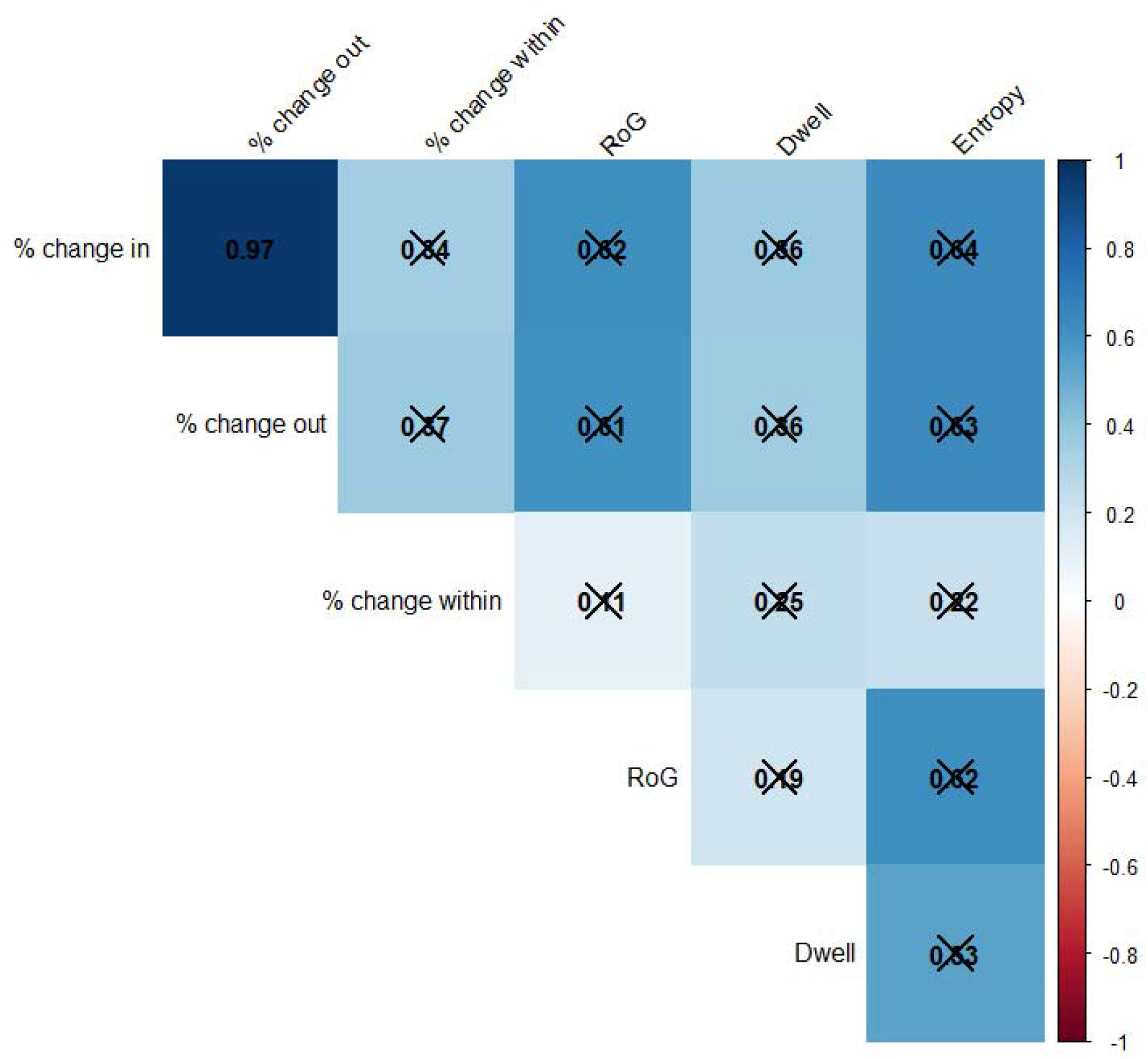

**Supp. Fig. 2.**
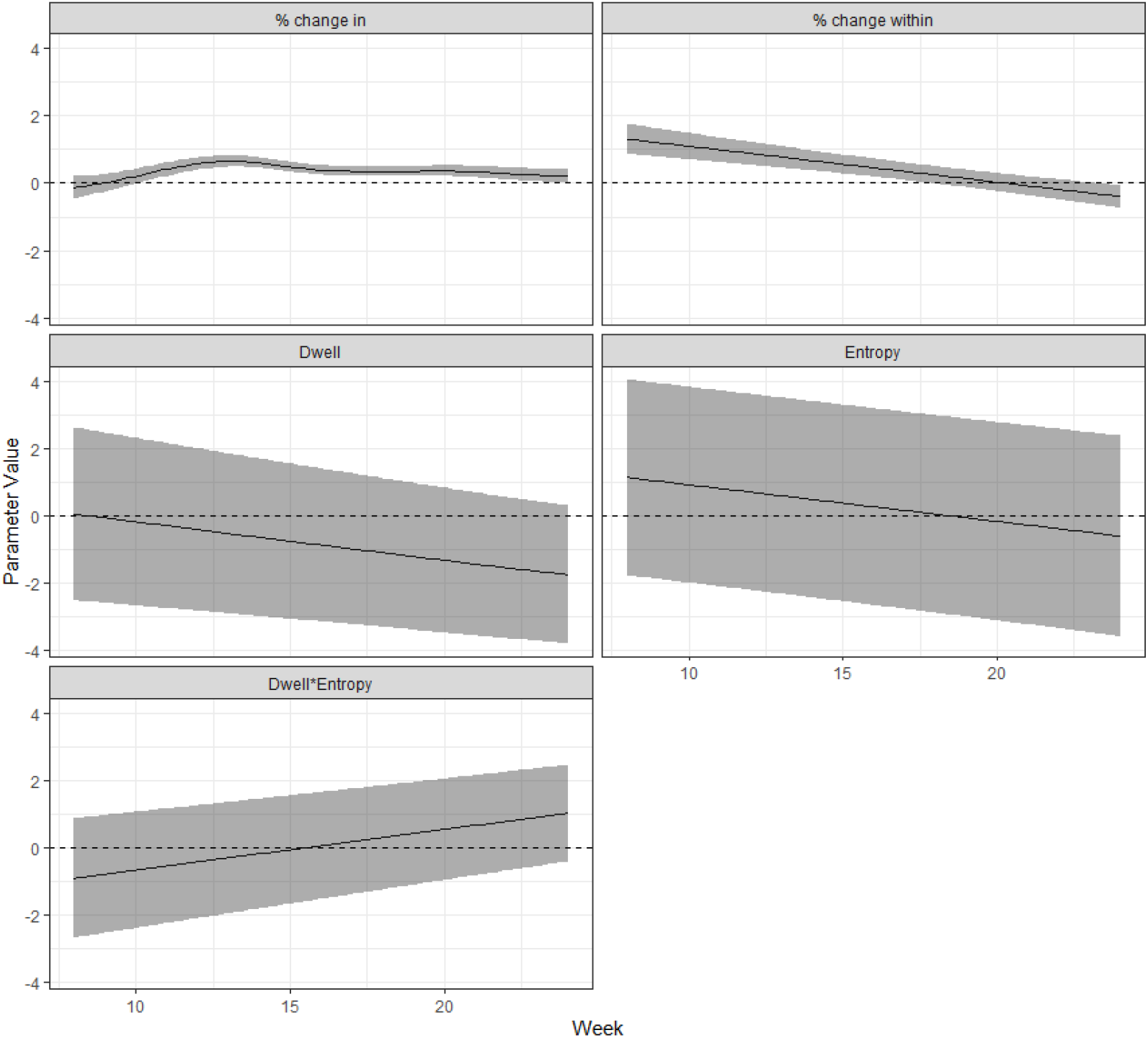

## Notes

### Competing Interest Statement

The authors have declared no competing interest.

### Funding Statement

No funding to report

### Author Declarations

Exemption provided by HSPH IRB.

